# Troponin HsTnI as pronostic factor in Covid-19 patients : Mohammed VI Hospital University experience

**DOI:** 10.1101/2025.05.16.25327634

**Authors:** Sabrina Belmahi, Oumaima Nassiri, Abdessamad Amrani, Amjad Idrissi, El-houcine Sebbar, Mohammed Choukri

**Affiliations:** Faculty of Medicine and Pharmacy of Oujda, Mohammed First University, Morocco; Central Laboratory, Mohammed VI University Hospital of Oujda, Morocco

**Keywords:** Troponin, Covid 19, Predective factor

## Abstract

**Background:** Recent reports have demonstrated high troponin levels in patients affected with COVID-19.

We sought in our study to determine the relationship between troponin levels and COVID-19 outcomes severity/mortality.

**Methods:** This was an observational retrospective cohort study of 325 patients admitted and diagnosed COVID-19 positive in Mohammed VI CHU in Oujda from July 2021 to November 2021

**Results:** The mean age between deceased and cured patients (68.92 ± 15.06 vs 62.5 ± 15.73 years, respectively, p<0.001) but no significant difference in the sex ratio was found in the two groups (p 0.379).

191 on admission, had a measured troponin ≤26 whom 80.6% were hospitalized in covid unit, while 134 patients had a measured troponin > 26ng/ ml whom 56.5% were admitted in the ICU (p<0.001)

73% of patients with an admission troponin ≤26ng/L survived while 27% had deceased with p<0.001.Similarly, the measurement of serum Troponin at the of hospitalization was 73ng/L (25.6–319.6) while for discharged patients was 9.6 ng/L (4.3–31.7)

The distribution of hs-TnI maximum value measured, showed a concentration significantly higher in the deceased patients vs survivors [200 ng/L IQR 48–651 vs 19 ng/L IQR 7.97–85.87, p<0.001].

**Conclusion:** our study demonstrates that Troponin may be an indicator of severity and may contribute to determining the severity of patients infected with COVID-19. Utilising troponin measurements alongside with other clinical and laboratory variables can be used for identifying patients at higher risk of in-hospital mortality and there by predict the progression of COVID-19 towards a worse outcome and to assign for more stringent strategies of treatment

## INTRODUCTION

COVID-19 is a new disease emerged in December 2019 in Wuhan, Hubei province China [1], This viral infection was rapidly spread to all countries and became the first large-scale pandemic of the 21st century. Patients with COVID-19 have developed several types of clinical manifestations, acute respiratory failure is the most common which have sometimes progressed to multiorgan failure or even death [2].

Social distancing measures including strict lockdowns imposed in many countries have successfully reduced the number of incident cases of COVID 19 infection; however, countries around the world have undergone successive waves of the pandemic in a scenario of alternate intensification and lifting of restrictions [3]. Vaccination campaigns are expected to minimize the risk of future waves and change the characteristics of hospitalized patients.

The list of complications of COVID-19 continues to grow longer every day, cardiac involvement seems to be among the most frequent after respiratory injury, major cardiac complications developed in a notable number of COVID-19 patients worsening the severe prognosis and possibly leading to death [4-5].

During the first wave of COVID-19, various retrospective studies of hospitalized patients provided a global view the determinants of severity and death [6-7]. Various Laboratory markers have been used for evaluating and predicting patients with severe COVID-19 disease [8]. Although variants of the virus have emerged with different severity and contagiousness these markers remain reliable and the evolution of treatment and a better knowledge of the virus has made it possible to reduce the severity of the disease [9-10].

We performed a study to assess whether the determination of high-sensitivity cardiac troponin I (hs-TnI) of patients hospitalized Mohammed VI University Hospital in Oujda (Reference hospital of the oriental region) in the second wave in Morocco is considered a prognostic factor in COVID-19 patients.

## MATERIALS AND METHODS

### 2.1 Study population and sample collection

Consecutive Covid-19 positive patients admitted to the Mohammed VI University Hospital in Oujda from July 2021 to November 2021 were included in this retrospective cohort study. A diagnosis of COVID-19 was based on SARS-CoV-2 real time reverse-transcriptase polymerase-chain-reaction (RT-qPCR) in respiratory samples performed in our laboratory, clinical, and radiological criteria according to in-house, national and international recommendations and guidelines.

#### Inclusion criteria

All those individuals who were tested positive for the SARS-CoV-2 according to the WHO and CDC guidelines for the detection and diagnosis of COVID-19 and who had at least one troponin assay.

#### Exclusion criteria

Patients who did not have a troponin measurement

### 2.2 Laboratory methods

#### 2.2.1 pre-Analytical

Blood collected on dry or heparinized tube. The collected samples were centrifuged at 4000 rpm for 10 minutes to obtain a clear supernatant serum.

#### 2.2.2 Analytical method

In this study, high sensitivity troponin I (hs-cTnI) was measured by using a high sensitivity chemiluminescent microparticle immunoassay (CMIA) using (ARCHITECT ci8200, Abbott Laboratories). Analytical method verification was performed; repeatability and intermediate precision were achieved using three levels of quality control. A method comparison was established between two Architect i-8200 automata. The results obtained show very satisfactory coefficient of variation according to norms and reference documents.

According to the manufacturer, the limit of blank is 0.7 to 1.3 ng/L, the 99th percentile concentrations are 34.2 ng/L for males and 15.6 ng/L for females, **and 26 ng**/ L Overall.

### 3. Statistical analysis

The statistical analysis of the data was performed using Statistical Package for the Social Sciences (SPSS) version21.0.Data were described as number and percentage, if categorical or mean ± standard deviation if continuous with distribution approximately symmetrical, or median and interquartile interval (IQR) if continuous with asymmetrical distribution or discrete. After checking for normality of distribution, differences between groups were explored with the student test or Mann–Whitney test and Ki2.

hs-TnI at admission was defined as the first measured troponin, hs-TnI near dead or at discharge was defined as the last measured troponine before dead or discharge.

## RESULTS

From 336 consecutive patients admitted to emergency department and tested for COVID-19, 325fulfilled the inclusion criteria and represented the present study population, 168(51.7%) were male individuals while 157(48.3%) were female individuals. Demographic characteristics and laboratory results of the study patients are summarized in Table 1.

**Table 1.** Clinical and demographic characteristics of 325 patients with COVID-19

|  | ALL | Deceased | Discharged | P Value |
| --- | --- | --- | --- | --- |
| **N %** | 325 | 111 | 214 |  |
| **Sex (M)** | 168 (51.7%) | 61 (36.3%) | 107(63.7%) | 0,379 |
| **(F)** | 157 (48.3%) | 51 (31.8%) | 107 (68.2%) |  |
| **Age years (mean ± SD)** | $64.7 \pm 15.760$ | $68 \pm 15.067$ | $62.54 \pm 15.715$ | <0,001 |
| **Trponin(first measurement) ((median, IQI)** | 20.60 (7.47–80.97) | 44.0 ng/L (21.8–164.00) | 12.0(5.20–37.40) | <0,001 |
| **Troponin, ng/L (max value during hospitalization) (median, IQI)** | 41.00 (11.00–240.00) | 200.00 (48.00–651.00) | 19.00(7.97–85.87) | <0.001 |
| **Troponin, ng/L (measurement before dead or discharge) (median, IQI)** | 20.00(6.90–108.00) | 73.00(25.60–319.60) | 9.60 (4.30–31.70) | <0.001 |

The mean age was 64.7 ± 15.76. Among them, 191 cases hospitalized in intensive care unit and 134 hospitalized in covid departments. The average length of hospitalization was 9 (6-15) days.

During the hospitalization, 111 patients died and 214 patients were treated and discharged.

There was a significant difference in mean age between deceased and cured patients (68.92 ± 15.06 vs 62.5 ± 15.73 years, respectively, p<0.001) but no significant difference in the sex ratio was found in the two groups (p0.379).

A total of 1156 hs-TnI measurements were performed during the hospital stay. Laboratory results showed median values of troponin atadmission in all included patients, of 20.6 ng/L [IQR 7.47–80.97] with concentrations markedly higher in the deceased patients vs survivors [44 ng/L IQR (21.8–164) vs 12 ng/L IQR (5.2–37.4), p<0.001].

We found that 73% of patients with an admission troponin ≤26ng/L survived while 27% had deceased. This difference was statistically significant p<0. 001.Similarly, the measurement of serum Troponin at the of hospitalization was 73ng/L (25.6–319.6) while for discharged patients was 9.6 ng/L (4.3–31.7)

The distribution of hs-TnI maximum value measured, showed a concentration significantly higher in the deceased patients vs survivors [200 ng/L IQR 48–651 vs 19 ng/L IQR 7.97– 85.87, p<0.001].

## DISCUSSION

Cardiac troponin I is a regulatory subunit of the troponin complex associated with the actin thin filament within cardiac muscle cells [11]. Troponin I, in conjunction with troponin C and troponin T, plays an integral role in the regulation of muscle contraction. Cardiac troponin is the preferred biomarker for the detection of myocardial injury based on improved sensitivity and superior tissues specificity compared to other available biomarkers of necrosis, including CK-MB, myoglobin, lactate dehydrogenase, and others [12]. Any elevation of troponin is synonymous with cardiac cellular suffering or lysis. When an increased value for cTnI is encountered in the absence of evidence of myocardial ischemia, a careful search of other possible etiologies for cardiac damage should be taken. Elevated troponin levels may be indicative of myocardial injury associated with heart failure, renal failure, chronic renal disease, myocarditis, arrhythmias, pulmonary embolism, or other clinical conditions. [13,14]

Although troponin values are usually high in chronic renal failure and elderly patients or elevated cardiac troponin values are frequent but the kinetics and high values in our series in patients with COVID-19, suggesting myocardial injury as a possible mechanism contributing to severe illness and significantly associated with fatal outcomes in some studies [15,16].

In the current study, patients who presented with High troponin level on their admission leading to their hospitalization in ICU. On the contrary, the lower the value of hs-TnI the lower the risk of admission to the ICU. Cohort studies from patients hospitalized with COVID-19 have shown that about 10–20% of patients had elevations in cardiac troponin, and that this was more common in patients admitted to the ICU [17,18,19, 20], which is coherent with our study. Therefore, initial measurement of hs-TnI immediately after hospitalization, followed by a longitudinal monitoring, can help clinicians to intercept dynamic changes in the release of hs-TnI as evidence of onset myocardial injury. In addition, increased levels of inflammatory biomarkers such as procalcitonin, ferritin, CRP, have been associated with severe COVID.

Several mechanisms may explain this phenomenon: viral myocarditis, cytokine-driven myocardial damage, microangiopathy, and unmasked CAD [21].Up to today, the exact etiology of heart damage and the main driver of troponin elevation in patients with COVID-19 remains under investigation [22]. The main pathophysiology of COVID-19 infection in severe cases may be related to the consequences of the cytokine storm. Therefore, it is possible to speculate that elevated levels of cardiac biomarkers in COVID-19 patients could be due also to a cellular damage related to wall stress caused by acute overload of the right ventricle. [5]

In discharged patients, we found in our study that the median Troponin was high on admission compared to discharge, unlike patients hospitalized in intensive care unit where the median Troponin increased from admission to discharge. Our series found that increased in troponine levels was associated with mortality and was significant. As reported in other studies such as Shi et al. [5]. Similar results were reported by Guo et al. [23], with highest mortality rates in those with elevated troponine levels.

Our results demonstrated that the serial Troponin measurement could predict the prognosis of Covid-19 patients, especially in patients with cardiac complications but our study has some limitations, first of all, this study was conducted in a single centre. However, the number of patients included is the highest compared to other published studies, and despite the high frequency of demand who had exposed in three times and the high cost of a troponin measurement, our laboratory was able to tackle the challenge.

## CONCLUSION

In conclusion, our study demonstrates that Troponin may be an indicator of severity and may contribute to determining the severity of patients infected with COVID-19. Our results should suggest to consider not only the respiratory symptoms, but also to triage patients with COVID-19 according to serially measurements of high-sensitivity troponin I, for identifying patients at higher risk of in-hospital mortality and there by predict the progression of COVID-19 towards a worse outcome and to assign for more stringent strategies of treatment.

## Data Availability

All data produced in the present work are contained in the manuscript

